# Implementing a Pharmacogenomic-driven Algorithm to Guide Antiplatelet Therapy among Caribbean Hispanics: A non-randomized prospective cohort study

**DOI:** 10.1101/2023.12.05.23299547

**Authors:** Héctor Nuñez-Medina, Mariangeli Monero, Lorna M Torres, Enrique Leal, Lorena González- Sepúlveda, Ángel M Mayor, Jessicca Y Renta, Edgardo R González-García, Ariel González, Kyle Melin, Stuart A Scott, Gualberto Ruaño, Dagmar F Hernandez-Suarez, Jorge Duconge

## Abstract

**Background:** After percutaneous coronary intervention (PCI), clopidogrel resistant patients are at an increased risk of major adverse cardiovascular and cerebrovascular events (MACCEs). We aimed to assess whether genotype-guided selection of oral antiplatelet drugs using a clinical decision support (CDS) algorithm reduces the occurrence of these ischemic events and improves outcomes among Caribbean Hispanic patients from Puerto Rico, who are underrepresented in clinical pharmacogenomic (PGx)-guided implementation studies.

**Methods:** Individual platelet function testing (PRU) measures, *CYP2C19*\*2 and *PON1* rs662 genotypes, clinical and demographic data from 8 medical facilities were included. Patients were separated into standard of care (SoC) and genotype-guided groups (150 each). Risk scores were calculated based on a previously developed CDS risk prediction algorithm designed to make actionable treatment recommendations for each patient. Alternative therapy with ticagrelor was recommended for patients with a high risk score ≥2. Statistical associations between patient time free of MACCEs and predictor variables (i.e., treatment groups, risk scores) were tested in this population using Kaplan-Meier survival analyses and Cox proportional-hazards regression models.

**Results:** Median age of participants is 67 years; BMI: 27.8; 48% women; 14% smokers; 59% with type-2 diabetes mellitus (T2DM). Among patients with high-risk scores who were free from MACCE events 6 months after coronary stenting, genotype-driven guidance of antiplatelet therapy showed superiority over SoC in terms of reducing the incidence rate of atherothrombotic events.

**Conclusions:** The clinical utility of our PGx-driven CDS algorithm to reduce the incidence rate of MACCEs among post-PCI Caribbean Hispanic patients on clopidogrel was externally demonstrated.

**Clinical Trial Registration Unique Identifier:** NCT03419325

## Introduction

Coronary heart disease (CHD) is the most common type of heart disease according to the Centers for Disease Control and Prevention (CDC), killing more than 382,000 people in 2020.^1^ According to the American Heart Association (AHA), Puerto Rico has the highest age-adjusted prevalence of CHD (6.6%) among all ethno-geographic regions.^2^ In the U.S., Hispanics >40 years old have more prevalence of cardiovascular risk factors for CHD such as high cholesterol, diabetes, high blood pressure, obesity, and smoking.^3–4^

Clopidogrel, a thienopyridine prodrug, is commonly prescribed as an antiplatelet therapy to prevent adverse cardiovascular outcomes (e.g., stent thrombosis) or reduce the risk of further ischemic events among patients undergoing percutaneous coronary interventions (PCI) for acute coronary syndromes (ACS) with ST-segment elevation myocardial infarction (STEMI), non-ST elevation myocardial infarction (NSTEMI) and unstable angina (UA); or among patients with cerebrovascular accident; stable coronary artery disease (CAD), specifically stable ischemic heart disease (SIHD); and peripheral arterial occlusive disease (PAD).^5–7^ The highly polymorphic CYP2C19-mediated pathway is the most important drug-metabolizing enzyme involved in the biotransformation of clopidogrel into an active metabolite that binds irreversibly to the ADP receptor P2Y12 on the platelet membrane to inhibit platelet aggregation.^8–9^ Despite its demonstrated efficacy, some studies have shown significant inter-individual variability in response due to *CYP2C19* genetic polymorphisms.^10–11^ The reduced function *CYP2C19*\*2 variant is strongly associated with a poor metabolizer phenotype, which produces less of the clopidogrel active metabolite and, hence, a poor clinical outcome.^12^ Patients who are resistant to clopidogrel are classified as having high on-treatment platelet reactivity (HTPR).^13^

According to the Updated Expert Consensus Statement on Platelet Function and Genetic Testing for Guiding P2Y12 Receptor Inhibitor Treatment in PCI, there is conflicting evidence on whether or not guiding treatment based on genetics is beneficiary.^14^ The POPular trial study reported a reduction of thrombotic events when using a genetic risk score-guided treatment in European PCI patients.^15^ However, Caribbean Hispanics are often underrepresented in pharmacogenomic studies, exacerbating healthcare disparities in this admixed and historically medically underserved population. Consequently, the clinical utility of implementing a genotype-guided antiplatelet therapy remains unclear for this population. This lack of information contributes to poor care management of this population and an increase in poor health outcomes.

The purpose of this study was to demonstrate the benefit of a pharmacogenetic-guided approach (PGx-CDS) over standard of care (SoC) in reducing the occurrence of major adverse cardiovascular and cerebrovascular events (MACCEs) among Caribbean Hispanic patients on clopidogrel. To reduce the incidence rate of MACCEs among Caribbean Hispanics on clopidogrel and validate a CDS tool designed to make actionable recommendations about the optimal treatment option in each patient, we aimed to assess statistical associations between patient time free of MACCEs and predictor variables (i.e., treatment groups, risk scores ≥2, T2DM) in this underrepresented population using Kaplan-Meier survival analyses and Cox proportional-hazards regression models.

## Methods

This was an open-label, multicenter, prospective, longitudinal, non-randomized clinical pharmacogenomic (cohort) study of antiplatelet treatment optimization conducted in patients undergoing PCI for ACS or SIHD and with documented extracardiac vascular disease (i.e., PAD) from the Commonwealth of Puerto Rico. The study was registered at ClinicalTrials.gov with unique identifier number: NCT03419325.

### Participants

A total of 300 Caribbean Hispanics patients on clopidogrel were recruited at eight medical facilities across the island of Puerto Rico (i.e., Cardiovascular Center of Puerto Rico and the Caribbean, San Francisco Hospital, Pavia Hospital, and UPR-Hospital Dr. Federico Trilla) from January 2018 through June 2022 and follow up over 6 months (final date of follow-up was December 2022). **Table 1** summarizes the inclusion/exclusion criteria for the study. All participants signed a written informed consent prior enrollment (approved IRB-protocol number A4070417). The **supplementary material** section provides further details on patients’ enrollment.

**Table 1.** Criteria for study inclusion and exclusion.

| Inclusion criteria | Exclusion criteria |
| --- | --- |
| Caribbean Hispanics residing in Puerto Rico<br>Both genders (i.e., males/females)<br>Age $\geq 21$ years old<br>Receiving clopidogrel (75mg/day) alone or in combination with aspirin as DAPT for therapeutic indications (ACS, stable CAD/ SIHD, PAD).<br>No clinically active hepatic abnormality.<br>The ability to understand the requirements of the study.<br>The ability to comply with the study procedures and protocols.<br>A female patient is eligible to enter the study if she is of childbearing potential and not pregnant or nursing, or not of childbearing potential. | Non-Hispanic patients<br>Currently enrolled in another active research protocols<br>BUN > 30 and creatinine > 2.0mg/dL<br>Platelet count <100,000/mm<sup>3</sup><br>Hematocrit (Hct) $\leq 25\%$<br>Nasogastric or enteral feedings<br>Acute illness (e.g., sepsis, infection, anemia)<br>HIV/AIDS, Hepatitis B patients<br>Alcoholism and drug abuse<br>Patients with any cognitive and mental health impairment<br>Sickle cell patients<br>Active malignancy |

### Collection of Specimens and Data

Individual clinical and demographic data were retrieved from electronic health records (EHRs). Up to two 3.0 ml 3.2% citrate tubes of whole blood were collected from each participant. For DNA isolation, 200 µL of blood were processed in the QIAcube following the QIAamp DNA Blood Mini Kit Protocol (QIAGEN, USA) and DNA quantification was performed using the NanoDrop 2000 Spectrophotometer.

### Platelet Functional Testing

Collected blood samples from each patient were used to individually determine residual on-treatment platelet reactivity with the VerifyNow® P2Y12 platelet function assay following manufacturer instructions. Results were reported as P2Y12 platelet reactivity units (PRU). A PRU cutoff value of 230 determined whether a patient was a poor (PRU ≥230; resistant) or normal (PRU <230; sensitive) responder to clopidogrel.

### Genotyping

TaqMan SNP genotyping assays were run to ascertain genotypes at two SNPs included in the CDS algorithm: *CYP2C19*\*2 (rs4244285) and *PON1* p.R192Q (rs662). StepOne™ Real-Time PCR System and software v2.3 were used to determine genotype calling following manufacturer instructions.

### Clinical Endpoints

The primary endpoint (MACCEs) was a composite of cardiovascular death, non-fatal myocardial infarction, ischemic stroke, definite/probable stent thrombosis, and hospitalization for revascularizations at 6 months after treatment. Reports of major and minor bleeding episodes were used as a secondary outcome. Major bleeding events included a need for blood transfusion, as well as any medical attention, admission to hospital, medical intervention or disability caused by the bleeding. In addition, minor bleeding events included bleeding gums, blood in urine, dark stools, bruising, nose bleeding, and blood vomiting. Adherence medication score was measured using the 4-item Morisky Medication Adherence Scale (MMAS-4).^16^

### Experimental Design

A full description of the study protocol has previously been reported.^17^ In brief, the CDS algorithm-guided actionable recommendations were implemented in the genotype-guided group, for patients with high-risk scores ≥2 to be escalated to ticagrelor; whereas those with low risk were kept or de-escalated to clopidogrel as shown in **Figure 1**. Recommendations for optimizing individual antiplatelet therapy were available through our custom mobile phone application (app) within 2 weeks of hospital discharge. To ensure full engagement, cardiologists were educated on proper use of the mobile app. This is a clinician-oriented, PGx-based point-of-care app that was developed in-house by our bioinformatics and technology (IT) team to allow the integration of patient’s genotypes into informed clinical decision making (i.e., CDS tool) so that clinicians may easily apply algorithmically guided DAPT plans in real-time, on-demand, in clinical settings. The following information was collected through record review: sex, age, BMI, co-medications, comorbidities, diagnosis of type 2 diabetes (T2DM), hematocrit (%), length and number of adjoined stents ≥30mm, left ventricular ejection fraction (LVEF, %), start of clopidogrel treatment, occurrence of MACCEs and time elapsed since starting therapy. Details on risk scores calculation and the PGx-CDS app used can be found in **supplementary materials** and **Figure S1**. In addition, flowcharts of clinical protocol in CAD/SIHD and patients with documented extracardiac vascular disease (i.e., PAD) or ACS patients undergoing PCI is depicted in **Figure S2A-B**.

**Figure 1.**
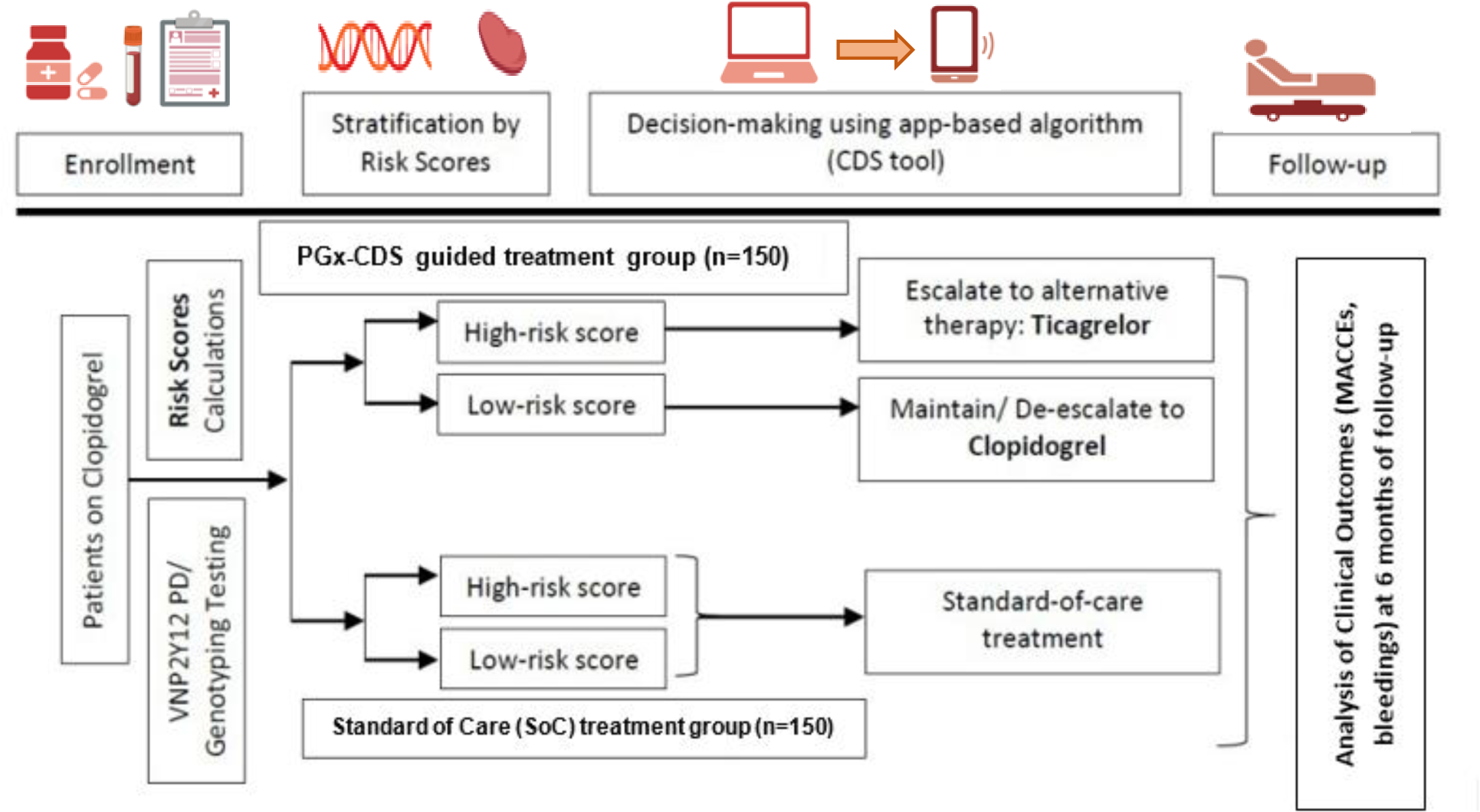
Experimental design. Matched non-concurrent sub-cohort of patients used as SoC controls. Data censored for 6 months. Further details can be found elsewhere.^17^

### Statistical Analysis

A study sample size of 140 patients per group was calculated to have 80% power to detect statistical differences with alpha of 0.05. To characterize the study cohort, a descriptive analysis of all demographics and clinical parameters was performed. Categorical data were summarized as frequencies and percentages. Continuous variables were reported as mean, ± standard deviation (SD) and standard error of mean (SE_M_). Statistically significant differences between groups were assessed, using either chi-square or Fisher’s exact probability tests for categorical variables, and two-tailed unpaired Student’s *t* test or Mann–Whitney U tests, as appropriate (i.e., when normality assumption was violated), for continuous variables (independent samples).

Cumulative survival (event-free time) curves for the occurrence of the major adverse endpoints during the follow up period were constructed using the Kaplan–Meier method, and differences were assessed using the log-rank test. Hazard ratios (HRs) with 95% CIs were obtained using Cox proportional hazards model. The length of survival (event-free time) was defined as the time of entry into the study until the occurrence of a major adverse event or termination of antiplatelet therapy. Termination of treatments were considered censored observations at the time of receiving their last pills. The Kaplan–Meier curves were truncated at 6 months of follow-up. In addition, a multivariate Cox proportional hazards regression analysis was performed to evaluate the effect of variables on the event-free time in these two groups. Selected variables included those showing significance in previous univariate analysis as well as others that were biologically or clinically relevant or that have been previously reported as significant in other studies. Adjusted and unadjusted HRs with 95% CIs were used to describe significant associations. All analyses were performed with STATA statistical software for data science (v18, StataCorp LLC, TX, USA).

## Results

**Table 2** summarizes baseline characteristics of participants in the study. The average age (median) of all participants in this study was 67 years old, with 52% self-identified as males. There was a high prevalence of conventional risk factors (i.e., high BMI of 27.8 kg/m^2^; 59% with T2DM diagnosis). Overall, 14% of participants were smokers, 88% were using statins (mainly simvastatin) to treat hyperlipidemia, 31% were on proton pump inhibitors (PPI, mainly pantoprazole) and 71% on aspirin as part of DAPT. Among participants, 72% had ACS and 18% had SIHD. Moreover, 9.7% reported the occurrence of MACCEs and 11% reported a major bleeding episode. Carriers of at least one *CYP2C19*\*2 allele represented 23% of all participants in both groups (MAF: 12.8%; 95%CI: 10.7-15.1), whereas carriers of the *PON1* rs662 (p.Q192R) variant represented about 60% of participants (MAF: 47%; 95%CI: 42.7-51.5). Of 150 patients assigned to genotype-guided therapy, 36 received a recommendation for switching to ticagrelor (24%, risk score ≥2). All participants assigned to the SoC received clopidogrel.

**Table 2.**
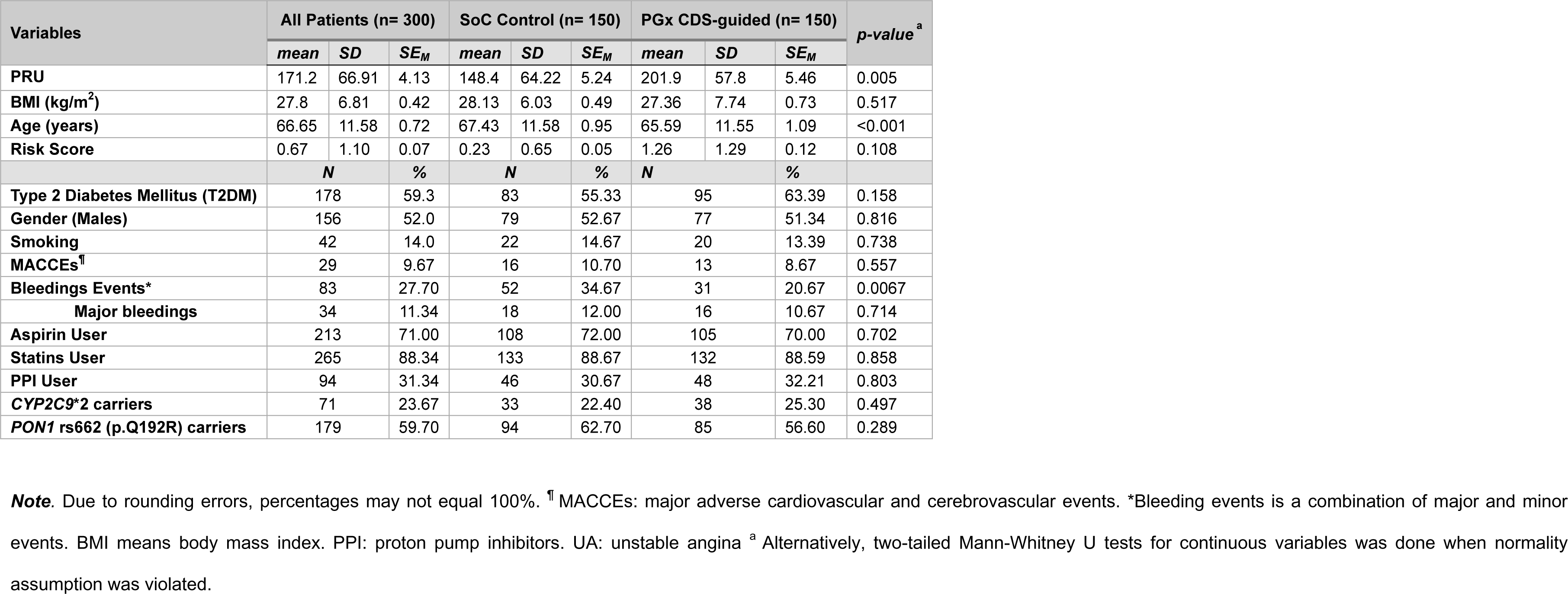
Baseline characteristics of study cohort participants (n=300, Caribbean Hispanics). Summary descriptive statistics and differences between treatment groups (SoC and Genotype-guided groups) were statistically tested by either two-tailed, unpaired *t*-tests for independent samples (continuous variables),^a^ or the chi-square/Fisher’s exact

Patient adherence scores above 82% were reported in both groups over the entire follow up period. The major reasons for non-adherence to treatment in this study were consistent with previously reported studies.^22–23^ In general, we observed high adherence scores for patients in both groups and for the whole period of assessment (i.e., 90%, 82% and 84% with high level of adherence after the first, third, and sixth month of follow-up, respectively). The most common reason for non-adherence to treatment was “*forgetting to take the medication*” (23%), followed by “*stopping taking the medication when they feel worse*” (9%) within the first six months of therapy. Other causes contributing to non-adherence were medication costs, transportation, and lack of communication with the cardiologist. Furthermore, no significant association was found between the levels of adherence and time of treatment (p=0.0570). Strategies to ensure medication adherence can help reduce worst outcomes.

Kaplan-Meier curves (survival analysis) and log-rank tests were used to assess the probability of participants being event-free after a follow up period of 6 months, when comparing MACCEs or bleeding occurrences over time between SoC and genotype-guided groups. The median follow-up period was 4.5 months. The occurrence of MACCEs at 6 months was reported in 13 and 16 patients in the genotype-guided and the SoC groups, respectively. Genotype-guided group had a lower but not significantly different risk of MACCEs (8.67% vs. 10.7%, hazard ratio [HR] = 0.56, 95% confidence interval [CI]: 0.19-1.56, p= 0.27) and bleeding (10.7% vs. 12.0%, HR = 0.52, 95% CI: 0.12-2.23, p= 0.38) compared with those in the SoC group. Among all Caribbean Hispanic patients who underwent PCI, genotype-driven CDS algorithm-guided selection of an oral P2Y12 inhibitor (i.e., clopidogrel vs ticagrelor), compared with SoC therapy, did not significantly reduce MACCEs occurrence over time based on the effect size that our study was powered to detect at 6 months. Our findings also showed that the overall frequency of MACCEs and major/minor bleeding episodes combined in the SoC was higher than that in the genotype-guided group (i.e., 40% and 23.2**%**, respectively). However, the incidence rate of these major adverse events (i.e., MACCEs and bleedings) was not significantly different between these two groups (i.e., 2.13 cases per 1,000 person-days versus 1.5 cases per 1,000 person-days, respectively; HR =0.67, 95% CI: 0.41-1.08, p=0.097) (**Figure 2**). Importantly, the unadjusted odds ratio (OR) for the association between these combinatorial adverse outcomes and treatment groups at 6 months was 2.205 (95%CI: 1.26 – 3.77, p= 0.0021), indicating that the odds of experiencing either a MACCE or a bleeding was two times higher among patients whose antiplatelet therapy was SoC versus those that were genotype-guided (**Figure 3**).

**Figure 2:**
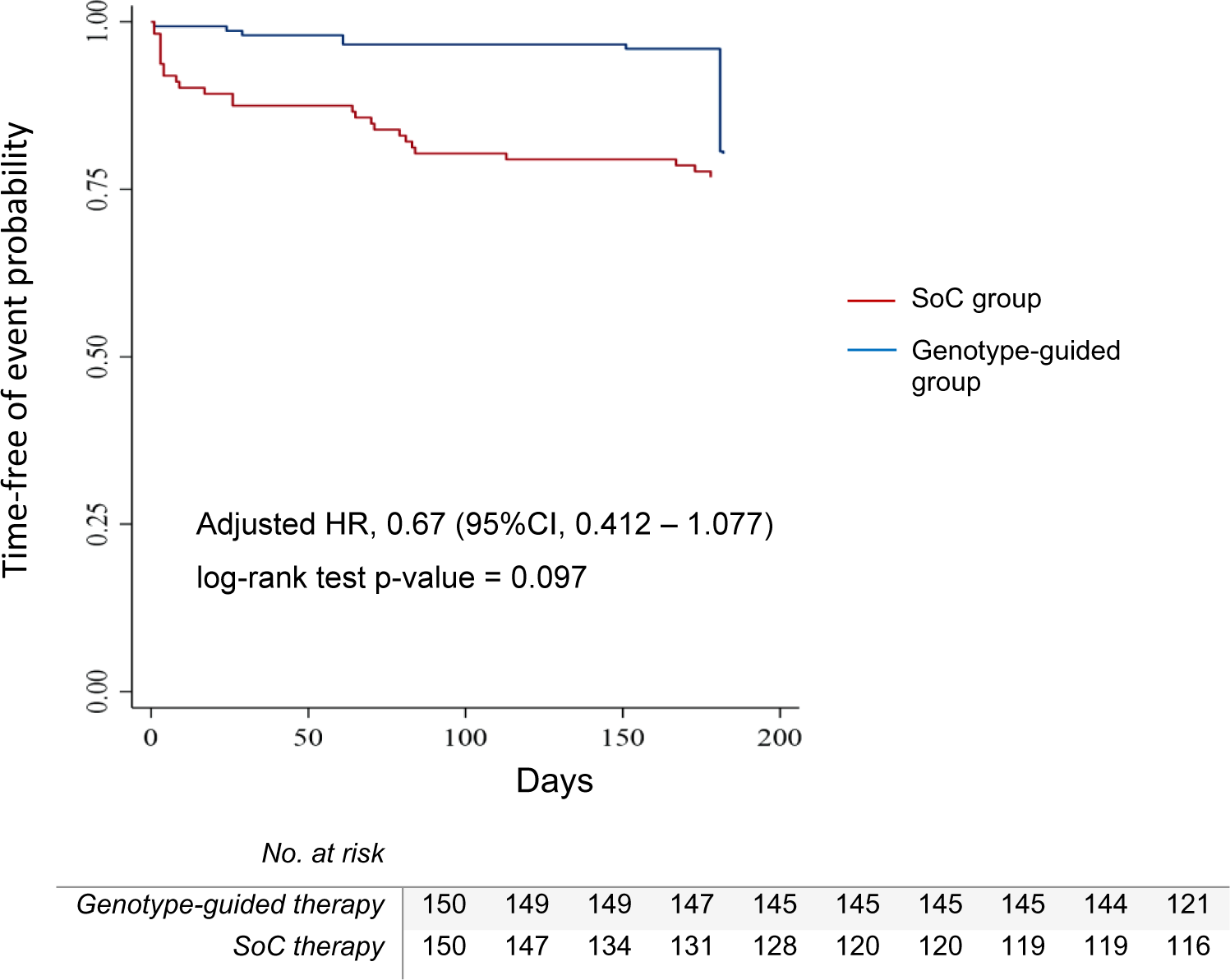
Kaplan-Meier plot of time free of MACCEs plus major bleeding episodes combined at 6 months of follow-up, grouped by treatment interventions (i.e., SoC vs. Genotype-guided groups). The adjusted hazard ratio (HR) and the corresponding log-rank test p-value are presented. Data censored for 6 months.

**Figure 3.**
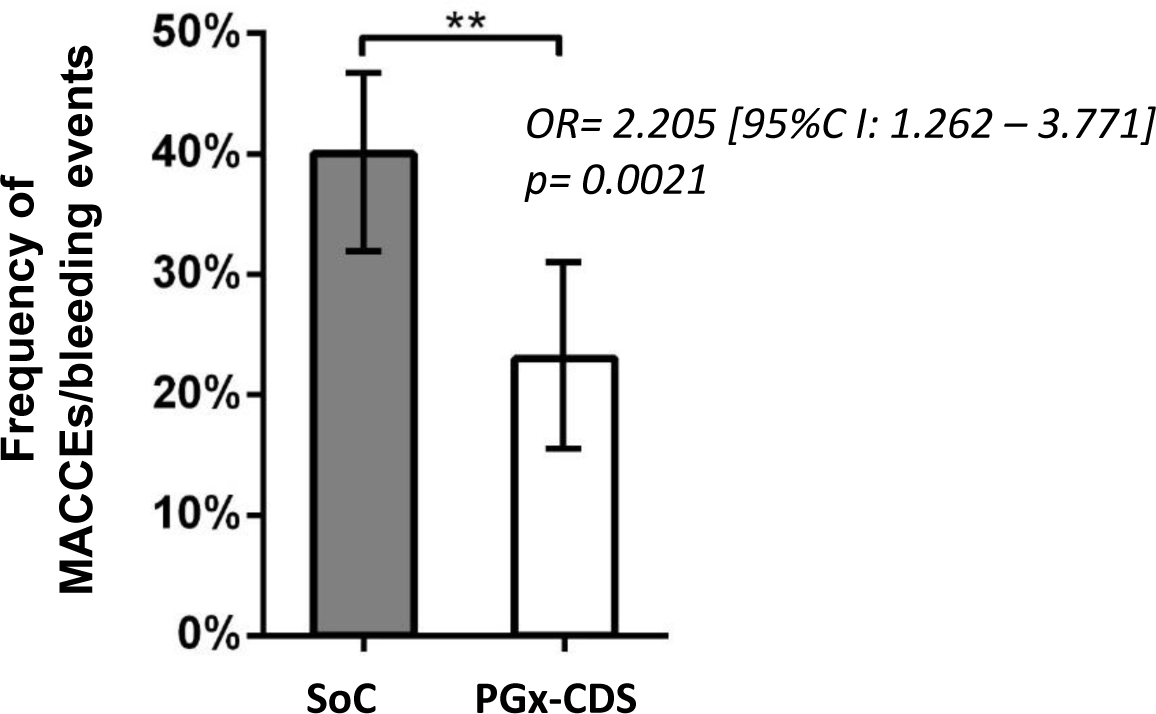
The PGx-guided CDS algorithm reduced the incidence rate of MACCEs and major/minor bleedings in Caribbean Hispanic patients. The incidence of MACCEs and major/minor bleeding events in the SoC group was 40% while in the genotype-guided group was 23%. The odds ratio (OR, 95%CI) and p-value of the association were computed using a one-sided chi-square test. Data are presented as frequency of events in patients.

Furthermore, when stratifying by patient risk score (i.e., high risk ≥2 vs. low <2), a significant difference in MACCEs occurrence at 6 months of follow-up was detected between the SoC and genotype-guided treatment groups among patients with high risk but not those with low-risk scores (**Figure 4**). Among high-risk patients, the incidence rate of MACCEs was 1.4 cases per 1,000 person-days in the genotype-guided group (8 out of 36 cases with events reported within a total person-time at risk of 5,772 days), whereas it was 11.39 cases per 1,000 person-days in the SoC group (11 of 14 cases with events reported in a total person-time at risk of 965 days). Escalation to ticagrelor in the genotype-guided group resulted in a significant reduction in the risk of MACCEs compared to the SoC group who continued receiving DAPT with clopidogrel (unadjusted HR: 0.15; 95%CI: 0.06-0.36). The log-rank test for equality showed a statistically significant difference in event-free time functions between the two groups (genotype-guided versus SoC, p<0.0001). Significant differences were also detected using multivariable Cox regression model, driven by BMI and T2DM covariates (adjusted HR: 0.104; 95%CI: 0.037-0.293; p< 0.0001). The forest plot in **Figure 5** shows the comparison of HRs calculated for the different analyses between study groups. Notably, the genotype-guided risk score discriminated between patients with and without high rates of MACCEs, indicating that this polygenic risk score is a useful predictor of poor clopidogrel outcomes among Caribbean Hispanics.

**Figure 4:**
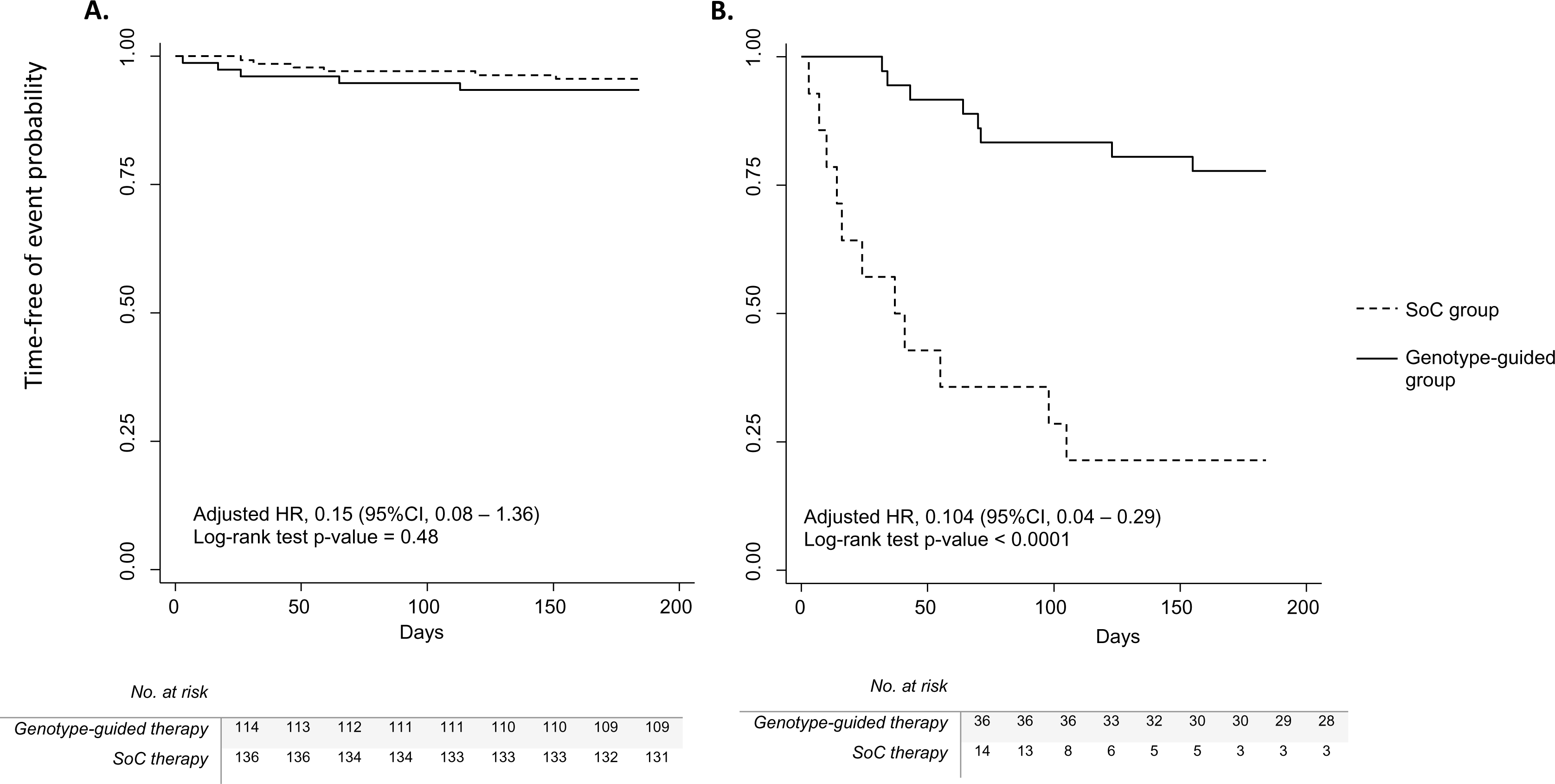
Kaplan-Meier plots of time free of MACCEs grouped by risk scores (6 months of follow-up). **Panel A** depicts the comparison between SoC and genotype-guided treatment groups for low-risk patients (risk score <2); whereas **Panel B** depicts the comparison between SoC and genotype-guided treatment groups for high-risk patients (risk score ≥2). Hazard ratios (HRs) and corresponding log-rank test p-values are shown, and data censored for 6 months.

**Figure 5.**
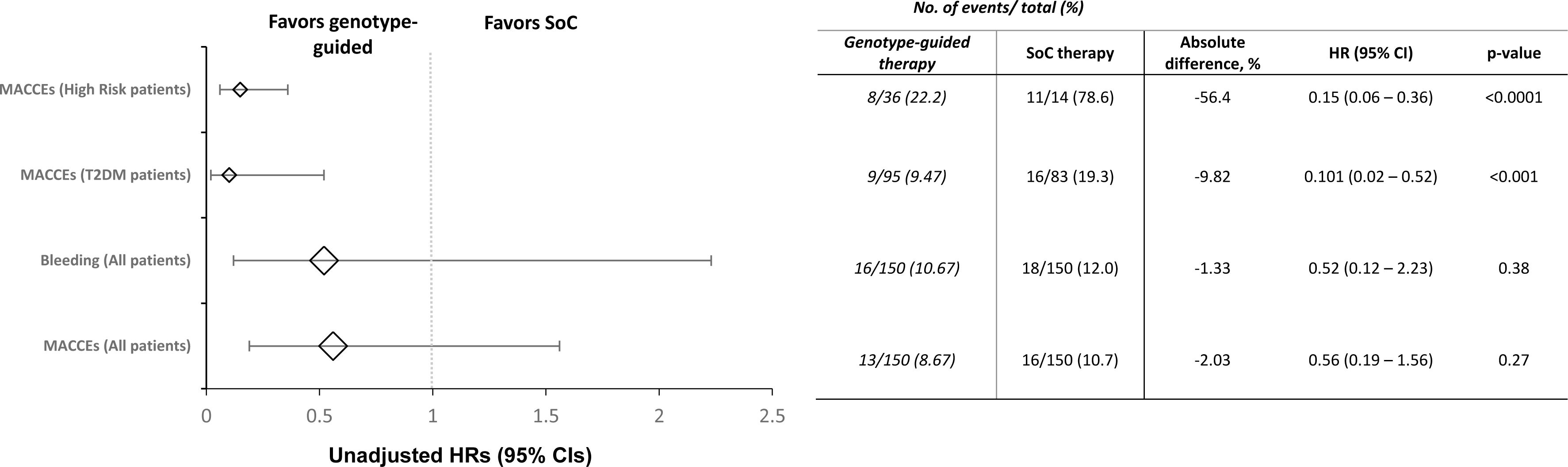
Forest plot of unadjusted hazard ratios (±95%CI) of MACCEs and bleeding for comparisons between treatment groups (i.e., PGx CDS vs SoC) and after stratifying by risk subgroups (high-risk) and relevant covariate (T2DM).

## Discussion

This clinical study aimed to investigate the efficacy of guiding anti-platelet therapy using a PGx-CDS algorithm in reducing the incidence rate of MACCEs among Caribbean Hispanics. Kaplan-Meier survival analyses and Cox proportional-hazards regression models were used to assess the time-to-event outcomes and identify potential risk factors associated with MACCE occurrence in this specific population. The findings from this research shed light on the impact of guiding anti-platelet therapy using a PGx-CDS algorithm in the context of Caribbean Hispanics’ cardiovascular health and provide valuable insights for future treatment strategies.

PGx-CDS has become a very useful tool to integrate individual genotyping information into existing clinical workflows and facilitate clinical translation of PGx. The use of our PGx-CDS platform as a mobile phone app to inform and guide cardiologists on DAPT prescribing decisions in real-time also represents a novelty in the incorporation of digital health among Caribbean Hispanics. Antiplatelet therapy switches were similar to those previously reported (i.e., 24% vs. 19%)^18^, with escalation to ticagrelor occurring mainly among patients with the nonfunctional *CYP2C19*\*2 allele. Notably, the occurrence of MACCEs was significantly lower in the genotype-guided group compared to the SoC group of our study cohort. MACCEs occurred 1.8-times less frequently in the genotype-guided group, with an incidence rate of 1.4 cases per 1,000 person-days versus 11.39 cases per 1,000 person-days in the SoC group, indicating a substantial risk reduction when guiding therapy by genetically driven risk status. Multivariable Cox model analysis also confirmed the significant differences and showed an adjusted HR of 0.104 (p=0.0001). These results suggest that the use of our risk score-based algorithm to individually guide antiplatelet treatment in Caribbean Hispanics offers superior outcomes in terms of reducing MACCEs in high-risk patients compared to the SoC approach. However, this benefit was not observed for patients at low risk or for the secondary outcome of bleeding episodes.

As shown in **Table 2**, baseline PRU was on-average significantly lower in the SoC than in the genotype-guided group (i.e., 148 vs. 202, respectively, p=0.005), which could have reduced the net effect of PGx-CDS guidance.

Of note, the thrombotic/bleeding trade-off of switching to ticagrelor/prasugrel versus continuing clopidogrel after PCI for high-risk patients remains unknown. In this study, significant differences in the occurrence of major/minor bleeding events were observed between the genotype guided and SoC groups (p=0.0067), but not between high (escalated to ticagrelor) and low (maintain/de-escalated to clopidogrel) risk patients (p=0.37). However, there was a trend in the expected direction of fewer reported bleeding episodes among patients who maintained or de-escalated to clopidogrel compared with patients who switched to ticagrelor. Published studies in other populations suggest that ticagrelor is associated with a higher risk of bleeding.^19–20^

The POPular Risk Score study reported a reduction of thrombotic events when applying a risk score guided treatment in patients who underwent PCI.^15^ Based on our findings, we conclude that the implementation of a PGx-guided CDS algorithm reduces MACCEs and improves health outcomes in Caribbean Hispanic CV patients undergoing PCI. Consequently, the adoption of a precision medicine paradigm that considers this genetic risk score-based strategy is expected to become a fundamental part of SoC when tailoring DAPT in this minority population with a high risk of thrombotic events. This study also contributes to address underrepresentation of minority populations in the field of clinical pharmacogenomics by being the first to validate a CDS tool for tailoring antiplatelet therapy in a Caribbean Hispanic population. Over the last decade, genotype-guided escalation, and de-escalation of antiplatelet therapy in post-PCI patients is becoming a standard practice in real-world clinical setting.^18,21^ Continuation of clopidogrel in high-risk patients is associated with adverse outcomes.

Kaplan-Meier survival analysis served as a crucial tool in this study to estimate the cumulative probability of MACCE over time among the Caribbean Hispanic cohort. The results revealed promising outcomes, indicating a significant reduction in MACCE among genotype-guided high-risk patients compared to the SoC. The survival curves displayed a clear separation between high-risk patients in the genotype-guided and SoC groups, supporting the favorable effect of guiding anti-platelet therapy by a PGx-CDS algorithm in mitigating adverse cardiovascular events in this population. However, this study also highlighted that subgroups within the population might experience varying benefits.

Cox proportional-hazards regression models were instrumental in identifying potential predictors of MACCE among Caribbean Hispanics on DAPT. Several demographic, clinical, and genetic factors were assessed, offering valuable insights into the multifaceted nature of MACCE occurrence risk in this specific population. The identification of these risk factors could aid in risk stratification and individualized treatment plans for Caribbean Hispanics, ultimately leading to improved patient outcomes. Further investigations are warranted to fully comprehend the underlying mechanisms and interactions between these risk factors to enhance the precision of preventive measures and therapeutic interventions.

Our study was not powered to detect significant differences among patients with and without the *CYP2C19*\*2 non-functional allele or the *PON1 rs662* (p.Q192R) variant. Nonetheless, among those carriers of at least one risk allele, a significant association with MACCEs was earlier found by our team (OR: 8.17, p=0.041).^30^ No significant effects were observed for the odds of having a single event in the study cohort. A similar *CYP2C19*\*2 MAF of 13.7% was reported in another cohort of Caribbean Hispanics.^29^ Our results also showed a *PON1 rs662* MAF of 47%, which is consistent with previous literature reports^24,27–28^. The inclusion of the *PON1* rs662 (p.Q192R) variant in our PGx-guided CDS algorithm is based on our prior findings in Caribbean Hispanics and those by others.^17, 24–26^ Therefore, this study supports in part the observed association of this variant with higher PON1 enzymatic activity and a lower risk of cardiovascular conditions.

In conclusion, this clinical study demonstrated that guiding individual anti-platelet therapy by novel a PGx-CDS algorithm is associated with a reduced incidence rate of MACCEs among high-risk Caribbean Hispanics (risk scores ≥2), as indicated by Kaplan-Meier survival analysis. The Cox proportional-hazards regression models provided important information on risk factors (i.e., T2DM, BMI, age), enabling a better understanding of cardiovascular risk in this population and guiding personalized treatment approaches. These findings contribute to the growing body of knowledge on the management of cardiovascular diseases in Caribbean Hispanics and underscore the importance of tailored interventions to optimize patient care and outcomes in this underrepresented population. Since age-adjusted cardiovascular mortality rate reduction across the US is lagging among populations experiencing significant health disparities as well as economic and social vulnerability, tailored interventions targeting Caribbean Hispanics are needed to close the gap.^31^ Nevertheless, the study’s limitations, including its moderate sample size, the use of a matched non-concurrent cohort of patients as SoC controls and potential selection bias, should be acknowledged, and further confirmatory study is warranted to corroborate and expand upon these findings.

## Data Availability

The data that support the findings of this study are available on request from the corresponding author (Dr. Jorge Duconge). The data are not publicly available due to restrictions as they contain information that could compromise the privacy of research participants.

## Acknowledgements

The authors thank the patients for voluntarily participating in this study protocol. A special acknowledgement to the Research Design and Biostatistics Core service of the Hispanic Alliance for Clinical and Translational Research (Alliance), for helping with study design, sample size calculations, and statistical analyses. Additionally, we also thank the Genomics Core of the CCRHD-RCMI Program at the UPR-MSC, for assistance with genotyping. Special thanks to Drs. Ednalise Santiago, Damian E. Grovas-Abad, Laura Ileana Fernandez-Morales, Luis Antonio Velez-Figueroa, Orlando Arce, Gretchen Gutiérrez, Mariela Loyola, Paola Pereira, as well as Frances Marín-Maldonado and Andrés López-Reyes for helping with sample and data collection.

## Author contributions

All the authors have accepted responsibility for the entire content of this research article and approved submission.

## Conflicts of Interest and Funding

The authors have no conflict of interest to declare. This work was supported in part by CCRHD-RCMI grant #2U54 MD007600 from the National Institute on Minority Health and Health Disparities (NIMHD) of the National Institutes of Health (NIH), the Postdoctoral Master in Clinical and Translational Research Program of the Hispanic Clinical and Translational Research Education and Career Development (HCTRECD) award (grant #R25 MD007607, NIMHD, NIH) and by the National Institute of General Medical Sciences (NIGMS)-Research Training Initiative for Student Enhancement (RISE) Program grant R25 GM061838. The Hispanic Alliance for Clinical and Translational Research (Alliance) is supported by the National Institute of General Medical Sciences (NIGMS), NIH, under award #U54 GM133807.

## Disclosures

The authors are solely responsible for the design and conduct of this study, all study analyses, the drafting and editing of the paper, and its final contents. The contents of this manuscript do not represent the views of the National Institutes of Health or the United States Government. No funded writing assistance was utilized in the production of this manuscript.

## Notes

### Competing Interest Statement

The authors have declared no competing interest.

### Clinical Trial

Clinical Trial Registration Unique Identifier: NCT03419325

### Author Declarations

The study secured Institutional Review Board (IRB) approval (# A4070417) and is therefore in compliance with institutional regulatory requirements. The IRB office at UPR-MSC (IORG000223; Federal-wise Assurance #FWA00005561) is approved by the Office for Human Research Protection (OHRP), Department of Health and Human Services.

xI have followed all appropriate research reporting guidelines, such as any relevant EQUATOR Network research reporting checklist(s) and other pertinent material, if applicable.

